# Reappraising Treatment Effect Heterogeneity in Schizophrenia: A Meta-analysis

**DOI:** 10.1101/2021.11.03.21265858

**Authors:** Robert A McCutcheon, Toby Pillinger, Orestis Efthimiou, Marta Maslej, Benoit H Mulsant, Allan H Young, Andrea Cipriani, Oliver D Howes

**Affiliations:** Institute of Psychiatry, Psychology and Neuroscience, Department of Psychosis Studies, King’s College of London, London, England; Psychiatric Imaging Group, MRC London Institute of Medical Sciences, Hammersmith Hospital, Imperial College London, London, UK; Institute of Clinical Sciences, Faculty of Medicine, Imperial College London, London, UK; Institute of Social and Preventive Medicine, University of Bern, Bern, Switzerland; The Krembil Centre for Neuroinformatics (KCNI), Centre for Addiction and Mental Health, Toronto, Ontario, Canada; Centre for Addiction and Mental Health, Toronto, Ontario, Canada; Department of Psychiatry, University of Toronto, Toronto, Ontario, Canada; Institute of Psychiatry, Psychology and Neuroscience, Department of Psychological Medicine, King’s College of London, London, England; Department of Psychiatry, University of Oxford, Oxford, UK; Oxford Health NHS Foundation Trust, Warneford Hospital, Oxford, UK; H Lundbeck A/s, 3 Abbey View, Everard Close, St Albans, AL1 2PS, UK

## Abstract

**Objective:** Determining whether individual patients differ in response to treatment (‘treatment effect heterogeneity’) is important as it is a prerequisite to developing personalised treatment approaches. Previous variability meta-analyses of response to antipsychotics in schizophrenia found no evidence for treatment effect heterogeneity. Conversely, individual patient data meta-analyses suggest treatment effect heterogeneity does exist. In the current paper we combine individual patient data with study level data to resolve this apparent contradiction and quantitively characterise antipsychotic treatment effect heterogeneity in schizophrenia.

**Method:** Individual patient data (IPD) was obtained from the Yale University Open Data Access (YODA) project. Clinical trials were identified in EMBASE, PsycInfo, and PubMed. Treatment effect heterogeneity was estimated from variability ratios derived from study-level data from 66 RCTs of antipsychotics in schizophrenia (N=17,202). This estimation required a correlation coefficient (*ρ*) between placebo response and treatment effects to be estimated. We estimated this from both study level estimates of the 66 trials, and individual patient data (N=560).

**Results:** Both individual patient (*ρ*=-0.32, p=0.002) and study level (*ρ*=-0.38, p<0.001) analyses yielded a negative correlation between placebo response and treatment effect. Using these estimates we found evidence of clinically significant treatment effect heterogeneity for total symptoms (our most conservative estimate was SD = 13.5 Positive and Negative Syndrome Scale (PANSS) points). Mean treatment effects were 8.6 points which, given the estimated SD, suggests the top quartile of patients experienced beneficial treatment effects of at least 17.7 PANSS points, while the bottom quartile received no benefit as compared to placebo.

**Conclusions:** We found evidence of clinically meaningful treatment effect heterogeneity for antipsychotic treatment of schizophrenia. This suggests efforts to personalise treatment have potential for success, and demonstrates that variability meta-analyses of RCTs need to account for relationships between placebo response and treatment effects.

## INTRODUCTION

Treatment effect heterogeneity refers to a situation in which the effects of a treatment differs between patients, for example some may receive marked benefit while others experience deleterious effects. Understanding the heterogeneity of treatment effects in addition to their mean magnitude, is important for clinical practice, but until recently it had not been meta-analytically investigated. Meta-analysis of variability has recently become an area of intense interest in psychiatric research. The technique has been applied to understand variability of brain structure (1), and subsequently brain function (2–5), immune function (6), and more recently to investigate variability in randomised controlled trials (RCTs) of therapeutic interventions thereby potentially providing an estimate of treatment effect heterogeneity (7–13).

The primary inferences drawn from studies of variability of clinical trial data relate to the presence or absence treatment effect heterogeneity, i.e. whether individual differences in treatment effects exist. A ‘treatment effect’ refers to the difference in the outcomes between placebo and active treatment that an individual would experience if all other circumstances were equivalent. It has been previously suggested that the existence of treatment effect heterogeneity would result in increased variability of symptomatic response to the active treatment arm, as compared to the placebo arm of an RCT (7; 9; 10). Several analyses of variability based on study-level data, however, found no evidence of greater variability in the active treatment arm, and thus concluded that treatment effects are likely to be relatively constant, which suggests the scope for personalisation of treatment is limited (7–13). These findings were surprising given the widespread clinical belief that patients differ substantially in their response to medication. These findings are also in contrast to previous research using individual patient data (IPD) that has suggested treatment effects vary between patients, with patients who are most severely ill at baseline benefitting the most from active treatment (14–17). While large IPD datasets provide a means to examine treatment effect heterogeneity, these are often not readily available. An advantage of variability meta-analyses is that only aggregate data is required, but the lack of consistent conclusions between the two approaches is concerning.

One explanation for these discrepant findings may be that the conclusions drawn from the variability meta-analyses rest on invalid assumptions regarding the correlation between the treatment effect and placebo response(11; 18). Specifically, the conclusions of these earlier meta-analyses are valid only when this correlation is zero or positive (Figures 1A, 1B). If the correlation is negative, treatment effect heterogeneity can exist even when the variability of the active treatment arm is no different to, or indeed of lesser magnitude than the variability of the placebo arm (Figure 1C) (10; 11; 18). However, this correlation between treatment effect and placebo response has not been previously estimated. As a result, formal estimation of the heterogeneity of treatment effects using aggregate data from RCTs has previously not been possible.

**Figure 1.**
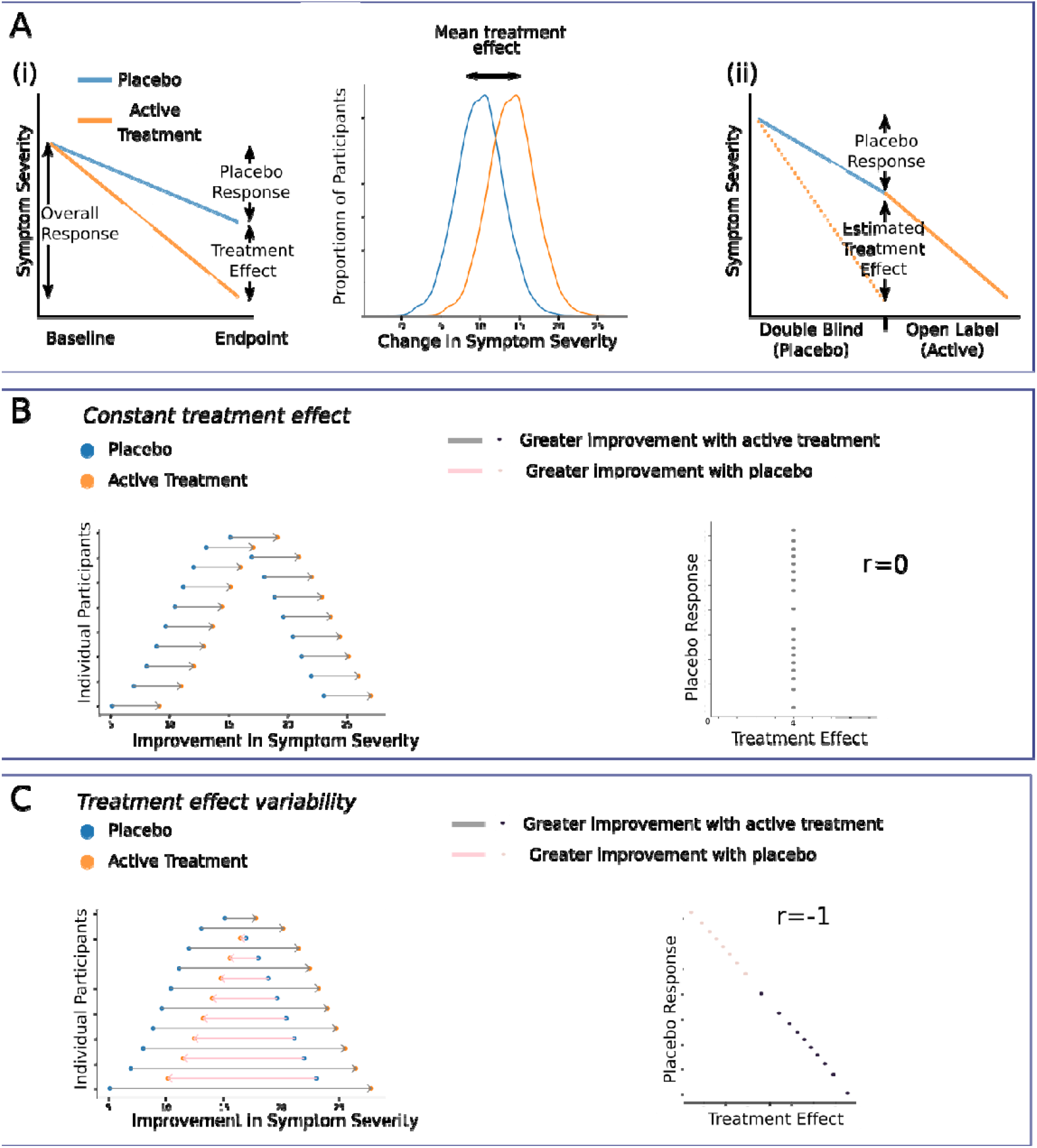
Inferring the variability of treatment effects A) (i) For a simple trial of parallel design, the average treatment effect can be estimated as the difference between mean outcome in placebo and mean outcome in active treatment. It is not possible, however, to directly determine the variability of individual treatment effects. (ii) Figure illustrates symptom change for a single participant initially receiving placebo followed by an open-label period of active treatment. Our ‘open label method’ estimation of treatment effects relies on the assumption that if this patient had taken drug in the double-blind period instead of placebo, the symptoms would change as indicated by the dashed line. B) The hypothetical case where for each patient we observe the outcomes in both treatment and placebo, and both have the same variability at the group level. All individuals display an identical treatment effect and there is therefore no correlation between placebo response and treatment effect. C) As in (B), both placebo and active treatment outcomes have the same variability at the group level. In this case, however, treatment effects and outcomes in placebo are negatively correlated at the patient level, and significant treatment effect heterogeneity exists.

It is therefore of major importance to estimate the correlation between treatment effect and placebo response, as a growing body of literature exists that cannot be accurately interpreted without this parameter. In the current paper, we estimate this value via complementary approaches, using both IPD and study level treatment effects from clinical trials. In conjunction with the results of a variability meta-analysis, this parameter allows for a formal estimation of the heterogeneity of antipsychotic treatment effects, as opposed to the primarily intuitive interpretations of previous meta-analyses(7; 9).

## METHODS

We searched for studies providing IPD from the Yale University Open Data Access (YODA) clinical trials database(19). We used this data to estimate the correlation between placebo and treatment effects at the individual patient level, using two alternative methods. We also used study level data from a recent meta-analysis of randomised controlled trials to estimate the correlation at the study level (7). We then used the most conservative of these coefficients to calculate the heterogeneity of treatment effects, using aggregate data from antipsychotic trials of schizophrenia(7). The estimates of treatment effect heterogeneity were then combined meta-analytically.

### Datasets

#### Individual Patient Data

All trials in the YODA database were searched to identify acute treatment clinical trials of antipsychotic medication in schizophrenia that included adults aged 18-65, who had a period of placebo treatment prior to a period of active treatment, with Positive and Negative Syndrome Scale (PANSS) scores recorded in both periods. Only a single trial met these criteria and it employed the following design. Individuals with schizophrenia who were receiving antipsychotic treatment and were symptomatically stable, were withdrawn from current medication and then randomised to placebo or active treatment for the duration of a 6-week double-blind period. Individuals who completed the double-blind period, or completed at least 21 days of double-blind treatment followed by discontinuation due to lack of efficacy, then entered an open-label extension where they received active treatment.

#### Study Level Data

We used all studies from a recent meta-analysis which had included all randomised double-blind placebo-controlled trials of antipsychotic monotherapy in the treatment of adults age 18-65 with schizophrenia (7). BPRS scores were converted to PANSS scores using the method described by Leucht et al. (20) to maximise the number of studies that could be included.

### Statistical Analysis

#### Estimating the correlation of treatment effect and placebo response from individual patient data

The average treatment effect for a population of interest is defined as the difference between the mean outcome if the whole population received placebo versus the outcome when receiving the active treatment. This can be estimated in RCTs (Figure 1). The treatment effect at the individual level cannot be estimated as easily as it refers to the difference between outcome under placebo and outcome under active treatment for the same patient with all other factors (including the moment in time) being identical. It is, however, under certain assumptions, possible to estimate a treatment effect for an individual. We performed this using two separate methods, which rest upon different assumptions.

The first method, termed subsequently the ‘open label method’ made use of one study from the YODA database, in which, for patients randomised to placebo, symptomatic change data was available for the same individual receiving both placebo and then active treatment. The placebo response for individuals randomised to the placebo arm during the double-blind period, was quantified as the change in PANSS score, between the start of the double-blind period and the point at which that individual left the double-blind portion of the trial. The estimated treatment effect was calculated as the change in symptom severity from the end of the double-blind period (during which the individual was receiving placebo) up until the timepoint that maximised the similarity between time spent in double-blind and open-label periods of the study (Figure 1A). This method relies on the assumption that the PANSS score we would observe if we could follow-up this patient under drug in the double-blind period (dashed line in Figure 1A; this is unobservable) equals the score we actually observe at the end of an-open label period of equal duration.

The second method, termed subsequently the ‘Linear Model (LM) method’, was based on a simple linear regression model. More specifically, we fit a linear model with symptom severity at the end of double-blind period as the outcome variable; and age, sex, and baseline severity entered as treatment modifiers (see supplementary). This method makes the usual assumption of linearity and additivity of the effects of the covariates and treatment on the outcome. Following model fitting, we were able to then estimate treatment effects at the individual level using that individual’s covariates.

The two methods described above allow us to estimate the individual treatment effects. Next, we estimated the Spearman correlation coefficient between placebo response and treatment effect for both methods, using only patients randomized to placebo. Note that placebo response is directly observable for these patients. This correlation was calculated for positive and negative subscales, and the total score.

In addition, for both methods we also calculated the correlation between overall response and both placebo response and treatment effect. These measures have relevance when considering how much information ‘overall response’ contains regarding treatment effects. This is relevant to attempts to develop predictive markers of treatment response, where initial studies are often undertaken in a naturalistic setting without a placebo arm, and use overall response as the outcome of interest.(21; 22)

#### Estimating the correlation of treatment effect and placebo response from study-level data

A Spearman correlation coefficient between study-level placebo response and treatment effect, weighted by the number of patients, was calculated using the package wCorr (Version 1.9.1) in R. This was performed for positive and negative symptom scales in addition to total symptoms scores. This method of calculating the correlation rests upon the assumption that the correlation between treatment effects and placebo response at the patient-level equals the one at the study-level.

Thus, we have described three alternative methods for calculating the correlation between placebo response and treatment effect. Below we describe how to use this *ρ* to calculate heterogeneity of treatment effects from aggregate data.

#### Estimating treatment effect heterogeneity

Due to a lack of previous estimates for the correlation between placebo and treatment effects, prior analyses of variability have not accurately quantified the magnitude of treatment effect heterogeneity (7; 9; 12; 23). They have instead reported a lack of difference between active and placebo response variabilities, that is a variability ratio (VR) that does not demonstrate a statistically significant difference from one. From this they have concluded that evidence for heterogeneity of treatment effects is lacking and the average treatment effect is a reasonable assumption for the individual patient.

*VR* is defined as follows, with *σ*_*AT*_ denoting the standard deviation of symptomatic change in the active treatment arm and *σ*_*PL*_ denoting the standard deviation of the placebo treatment arm:

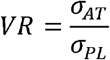

The variable of clinical interest, however, is the standard deviation of the treatment effect (*σ*_*TE*_). This can be estimated if *VR* and the correlation (*ρ*) between placebo response and treatment-effect are known (see (11; 18) for further description):

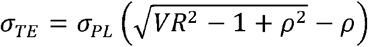

Note that for *VR*^2^ < 1 there is also an alternative solution for *σ*_*TE*_; see the Appendix and also(11; 18) for details. After estimating *ρ* as described previously (using whichever of the methods gave the most conservative value, i.e. the value corresponding to the correlation of lowest absolute magnitude), we next calculated VR using the published study-level data for RCTs of antipsychotic treatment of schizophrenia. Subsequently we estimated *σ*_*TE*_ using the formula above. Finally, we combined the values of *σ*_*TE*_ from all studies via a random effects meta-analysis implemented by the package ‘meta’ (version 4.18-2) for R (version 3.6.3) with between study inconsistency quantified using the I^2^ statistic(24; 25). In addition to a single summary estimate across all trials meta-analyses were also performed with studies grouped according to antipsychotics used. Meta-analyses were also performed for positive and negative PANSS subscales where these were reported, using the relevant value of *ρ* calculated above. In a sensitivity analysis we redid the calculations using instead the most liberal value (i.e. the value corresponding to the correlation of highest absolute magnitude) for *ρ* among the three methods.

To put the estimate of variability into perspective and to help assess its clinical importance, we also estimated the average treatment effect in the same RCTs. More specifically, we performed a random effects meta-analysis using the observed mean difference between drug and placebo arms.

Reported p values are two sided, and test against a null hypothesis of both the correlation (*ρ*) and the heterogeneity of treatment effects not differing from zero.

## RESULTS

### Individual Patient Data

Of the 68 studies examining antipsychotics in schizophrenia, 1 trial was eligible (clinicaltrials.gov registration: 00650793, see Table 1). This included 88 individuals who received placebo treatment during the double-blind period, and 384 individuals who received antipsychotic treatment during the double-blind period. Full details of the trial are available at https://yoda.yale.edu/.

**Table 1.** Characteristics of the clinical trial providing individual patient data Participant details reflect those who completed the double-blind phase of the trial and subsequently entered open-label phase, i.e. those participants included in the current analysis. The duration of open-label column refers to the duration up until the final symptom rating was taken for the purposes of the current analysis (i.e. to match the double-blind period as closely as possible).

| YODA number | Clinical trial number | Description | Double-Blind Treatment Drug | Open-Label Treatment Drug | N | Age, Years Mean (SD) | Sex % female | Duration double blind, Days mean (SD) | Duration open label, Days Mean (SD) |
| --- | --- | --- | --- | --- | --- | --- | --- | --- | --- |
| Sch-703 | 00650793 | 6-week double-blind period followed by 52-week open-label period | Placebo | Paliperidone (3-12 mg daily) | 88 | 38.1 (10.6) | 51.1 | 35.7 (10.0) | 33.8 (16.9) |
|  |  |  | Paliperidone (6-12mg daily),<br>Olanzapine (10mg daily) |  | 384 | 36.6 (10.8) | 50.5 | 40.8(6.2) | 37.3(15.1) |

The open label method yielded a strong negative correlation between the estimated treatment effect and placebo response for PANSS total (ρ=-0.62, p <0.001), positive (*ρ*=-0.61, p<0.001), and negative (*ρ*=-0.35, p<0.001) scales (Figure 2A). The assumption underlying the open label method is supported by the fact that PANSS scores at the end of the double-blind period for those initially randomized to antipsychotic treatment were similar to the scores at the end of the open label period for those initially receiving placebo (see supplementary eFigure 1). A strong positive correlation was observed for overall response and placebo response (*ρ*=0.68, p <0.001), while no correlation was observed between overall response and treatment effect (*ρ*=0.05, p=0.64).

**Figure 2.**
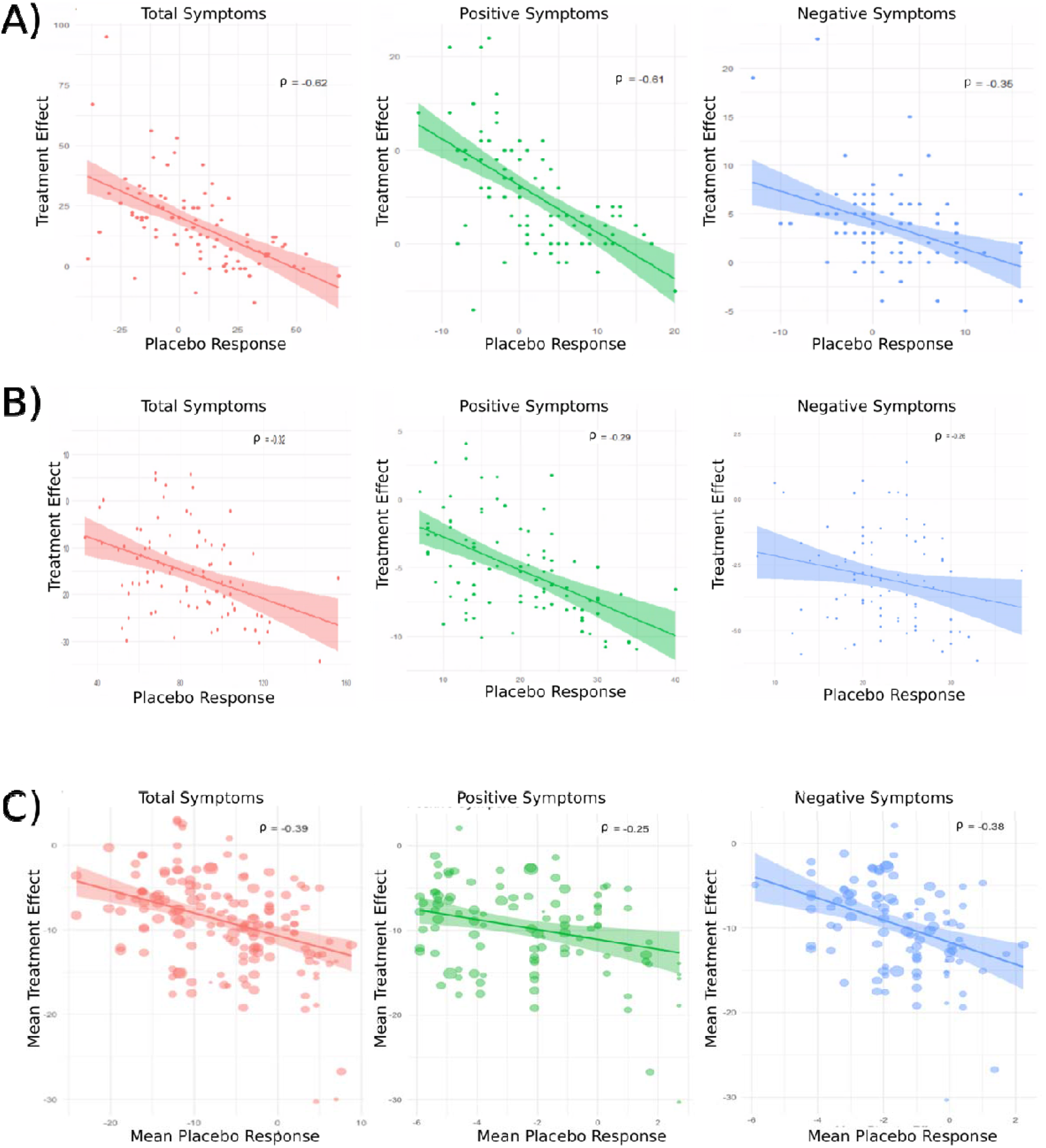
Correlation between treatment effects and placebo response A negative correlation between treatment effect and placebo response is observed for total, positive, and negative scales using all three approaches. A) Correlation between treatment effects estimated using the open label method and observed placebo response. Each point represents a participant. B) Correlation between treatment effects estimated using the LM method and observed placebo response. Each point represents a participant. C) Relationship between study level estimates of treatment effect and placebo response. Each point represents a clinical trial, with the size of the point proportional to the number of subjects in the trial.

The LM method also yielded a negative correlation between the estimated treatment effect and placebo response, this was observed for PANSS total (*ρ*=-0.32, p=0.002), positive (*ρ*=-0.29, p=0.006), and negative (*ρ*=-0.26, p<0.013) scales (Figure 2B). A strong positive correlation as observed for overall response and placebo response (*ρ*=0.90, p <0.001), while no correlation was observed between overall response and treatment effect (*ρ*=0.07, p=0.50).

Both methods estimate individual treatment effects and thereby allow for estimation of treatment effect heterogeneity within the trial. Clinically meaningful heterogeneity was apparent with both methods when observing the distribution of treatment effects (see supplementary eFigure2).

### Study Level Data

Data were analysed from 66 clinical trials including 17,202 patients (See supplementary for PRISMA flow diagram and checklist) (7). Across studies, we found a moderate negative correlation between placebo response and treatment effect for total (*ρ*=-0.39, p<0.001), positive (*ρ*=-0.25, p=0.01), and negative (*ρ* =-0.38, p<0.001) symptoms (Figure 2C).

Thus, we concluded that all three methods (which employ different assumptions) gave consistent evidence of negative correlation between placebo response and treatment effect.

### Treatment effect heterogeneity from study level data

Using the LM method values of *ρ*, as these were the most conservative, a meta-analysis of treatment effect heterogeneity estimated standard deviation for total symptoms to be 13.5 PANSS points (95% Confidence Interval (CI) 12.7 to 14.3, p<0.001, I^2^=45%) (Figure 3). For positive symptoms the estimate was 4.0 (95%CI 3.7 to 4.3, p<0.001, I^2^=53%), and for negative symptoms the estimate was 2.8 (95%CI 2.5 to 3.1, p<0.001, I^2^=57%) (Figure 3).

**Figure 3.**
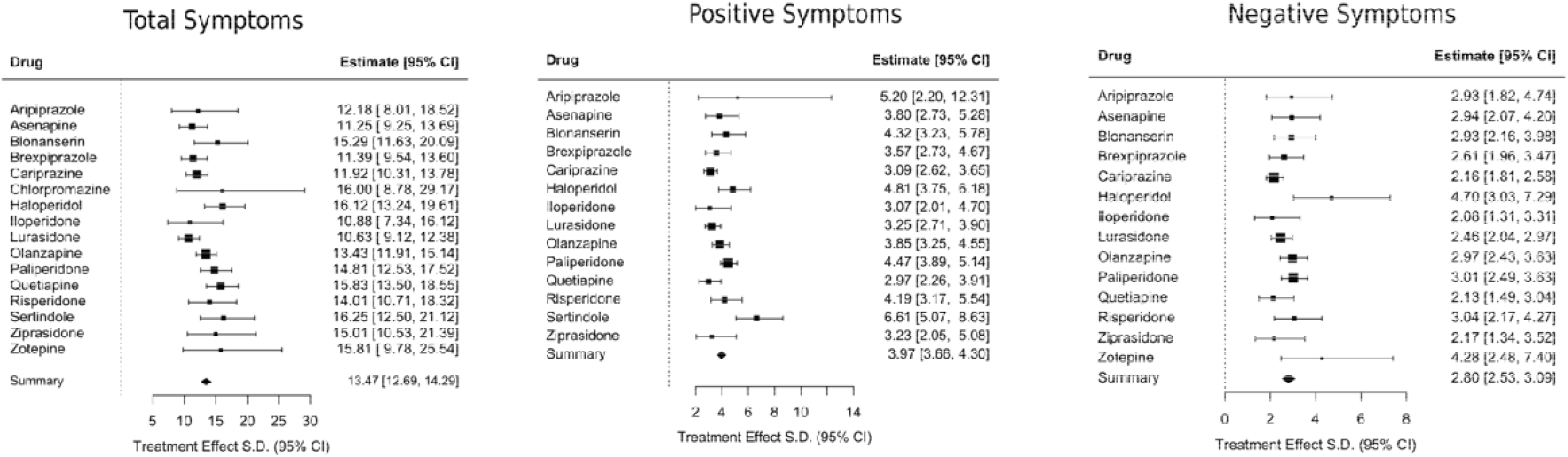
Meta-analysis of within-study estimates of the standard deviation (SD) of patient-level treatment effects of antipsychotics versus placebo. Results are shown by drug as well as in a summary, i.e. across all drugs.

The mean treatment effect was estimated as 8.6 (95% CI 7.8 to 9.4, I^2^=38%, p<0.001) for total symptoms, 2.7 (95%CI 2.3 to 3.1, I^2^=35%, p<0.001) for positive symptoms, and 1.8 (95%CI 1.5 to 2.0, I^2^=27%, p<0.001) for negative symptoms.

The expected distribution of treatment effects based on these values is illustrated in Figure 4. For total symptoms this distribution equates to an individual located at the 75^th^ centile benefiting from a treatment effect of a 17.7 point improvement on the PANSS, compared to an individual at the 25th centile receiving a worsening of symptoms of 0.5 points compared to placebo. For positive symptoms the 75^th^ centile equates to an improvement of 5.4 points, and the 25^th^ centile equates to a worsening of 0.05 points. For negative symptoms the 75^th^ centile equates to an improvement of 3.7 points, while the 25^th^ centile equates to a deterioration of 0.1 points. The distribution suggests that 74%, 75% and 74% of patients will show a non-zero benefit in terms of total, positive symptoms, and negative symptoms respectively.

**Figure 4.**
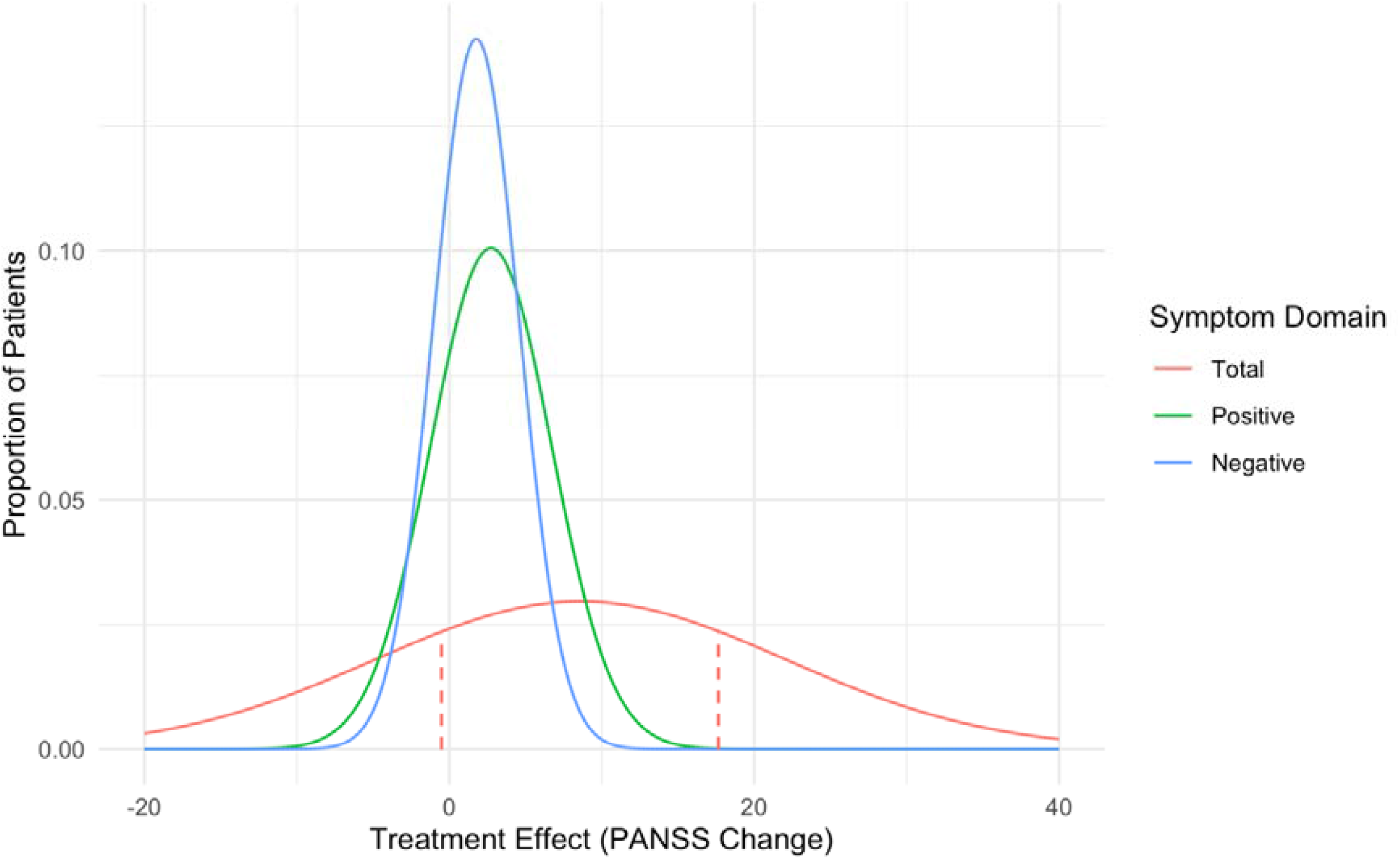
Treatment effect variability in antipsychotic treatment Clinically meaningful variability in treatment effects of drug versus placebo is apparent for total, negative, and positive symptoms. Dashed lines represent upper and lower quartiles for total symptoms. A positive treatment effect favours drug over placebo.

Finally, when we used the least conservative estimates for *ρ* the distribution of treatment effects was wider, pointing to even larger variability of treatment effects (see supplementary).

## Discussion

We found a negative correlation coefficient between placebo response and treatment effects after employing three different estimation methods, using both individual-as well as study-level data. This finding, when combined with the variability ratio observed in clinical trials, implies the existence of marked heterogeneity of patient-level treatment effects. Our more conservative estimates showed that a quarter of individuals experience a substantial benefit of over 17 PANSS points and a quarter experience a deleterious effect relative to placebo. Clinically important treatment effect heterogeneity was also estimated for positive and negative symptom domains. This finding was also corroborated when directly modelling the patient-level treatment effect using the IPD we had available.

On the basis that individual patients differ from one another in terms of overall response to antipsychotic treatment, it has long been assumed that they also differ in terms of the benefit they get from the drug. This has been supported by findings from individual patient data meta-analyses (14–16). This interpretation was recently challenged by meta-analyses which suggested that antipsychotics may in fact deliver a relatively constant treatment effect, and that clinically observed variability was therefore secondary to variability of placebo response, treatment-unrelated fluctuation in symptom severity, and measurement error (7; 9). In the current paper we bridge the gap between the conclusions of variability meta-analyses and those of IPD analyses and clinical experience. We show the evidence is consistent, with the discrepancy in conclusions resulting from a previously imprecise interpretation of the variability ratio. Specifically, previous meta-analyses of variability did not formally tie the variability ratio to the outcome of interest: the heterogeneity of treatment effects. The current analysis undertakes this vital step, and as a result demonstrates that the findings of variability meta-analyses, individual patient data meta-analyses, and clinical experience are consistent, and point to the existence of meaningful heterogeneity of antipsychotic treatment effects in adult patients with schizophrenia.

### Limitations

Our open-label method for estimating individual treatment effects involved calculating placebo responses in individuals who had previously received antipsychotic treatment. This is unavoidable due to a lack of available trials of suitable design in antipsychotic-naïve individuals but has potential disadvantages. In addition to possible carryover effects, withdrawal effects and placebo responses are enmeshed, and as a result our estimates of variability may partly reflect the variability of withdrawal effects. To disentangle withdrawal and placebo responses in crossover designs is complex but not impossible, and could be considered in studies aiming to further unpack individual variability of response (26). The open label method also assumes that the change in symptom severity with an active compound following a period of placebo treatment is a fair estimate of the treatment effect; whether this is fully justified is not known, although the group level findings suggest this may be a reasonable assumption.

The LM method for estimating treatment effects does not employ data from the open label phase and so does not rely on the same assumptions. It does however estimate treatment effects and placebo response after assuming linear, non-interacting relations between symptom scores and the baseline covariates of age, sex, and symptom severity. Moreover, given that other covariates are likely to play a significant role in determining both placebo response and treatment effects the estimates produced may not be entirely accurate.

The study level calculation of the placebo-treatment effect correlation circumvents some of the shortcomings of the individual level analyses. This analysis, however, runs the risk of aggregation bias (“ecological fallacy”), i.e. a correlation observed across studies at the study level may not reflect correlation at the individual level.

Concerns about the three approaches are mitigated by the consistency of findings between them. In addition, a negative correlation was a priori expected, as the greater a placebo response in an individual the less room there is for an additional treatment effect, and there are not reasons to believe that any of the methods would produce bias towards a negative correlation. In addition, we believe that our estimate of the correlation coefficient is the best available current estimate and as such its use is indicated, as the use of some form of coefficient is required to make any form of valid inference from variability ratios.

### Future work

The current study provides empirical support for the hypothesis that interindividual heterogeneity in treatment effects exists. The correlation coefficient between placebo and treatment effects, a central part of the analysis, was, in the case of the individual patient data, calculated from a clinical trial of paliperidone. Future work examining other medications is necessary, to investigate whether this coefficient differs between drugs. This will allow determination of whether antipsychotics differ in terms of treatment effect heterogeneity. It will also be of interest to see if outcome measures other than symptom rating scales, such as functioning, adverse effects, and cognitive measures show similar heterogeneity of treatment effects.

Much of the work aimed at identifying markers for treatment response uses overall response as the outcome of interest (21; 22). However, overall response comprises two components - a treatment effect and a placebo response. We found that while strongly related to placebo response, overall response showed almost no correlation with treatment effect. As a result, markers of overall response identified in this fashion may be of benefit in identifying patients who are likely to improve but they may fail to identify those who will most benefit from treatment. Future work may therefore benefit from using measures of treatment effect rather than overall response as the outcome of interest. The treatment effect is more difficult to estimate than a measure of overall response but it can be done using data from either sufficiently large parallel group RCTs with adequate measurement of relevant covariates, or from trials employing cross-over designs.

Other psychiatric treatments including antidepressants (8; 13), and brain stimulation (12) have also recently been examined in meta-analyses of variability ratios. As with the initial variability analyses of antipsychotic trials, the initial conclusion of these studies has been that minimal interindividual heterogeneity of treatment effects exists. However, these conclusions depend on the assumption of a positive correlation between placebo and treatment effects, which as the results presented above demonstrate, may well not be the case. Future work should seek to identify the correlation between treatment effects and placebo responses in other disorders and with other treatments. The estimation of this correlation will then allow for determination as to whether interindividual heterogeneity of treatment effects also exists in these disorders.

## Conclusion

The current findings support the hypothesis that substantial interindividual heterogeneity exists in terms of symptomatic response to antipsychotic treatment in schizophrenia. In turn, these findings support efforts to provide treatment personalisation. Future work should aim to identify which medications and symptom domains are most likely to benefit from personalised precision approaches, and aim to identify predictors of treatment effect in addition to predictors of overall response.

## Data Availability

All data are available at https://pubmed.ncbi.nlm.nih.gov/ and https://yoda.yale.edu/

## Disclosures

Dr McCutcheon has received honoraria for educational talks from Otsuka.

Dr Pillinger has participated in speaker meetings organised by Lundbeck, Otsuka, Sunovion, Schwabe Pharma and Recordati.

Dr. Mulsant receives research support from Capital Solution Design LLC (software used in a study founded by CAMH Foundation), and HAPPYneuron (software used in a study founded by Brain Canada). Within the past five years, he has also received research support from Eli Lilly (medications for a NIH-funded clinical trial) and Pfizer (medications for a NIH-funded clinical trial). He has been an unpaid consultant to Myriad Neuroscience Professor Cipriani has received research and consultancy fees from INCiPiT (Italian Network for Paediatric Trials), CARIPLO Foundation and Angelini Pharma, outside the submitted work.

Professor Howes is a part-time employee of H Lundbeck A/s and has received investigator-initiated research funding from and/or participated in advisory/ speaker meetings organised by Angellini, Autifony, Biogen, Boehringer-Ingelheim, Eli Lilly, Heptares, Global Medical Education, Invicro, Jansenn, Lundbeck, Neurocrine, Otsuka, Sunovion, Rand, Recordati, Roche and Viatris/Mylan. Neither Professor Howes nor his family have holdings or a financial stake in any pharmaceutical company. Professor Howes has a patent for the use of dopaminergic imaging.

## Acknowledgements

Dr McCutcheon is supported by the National Institute for Health Research (NIHR).

Dr Pillinger is supported by the NIHR and Maudsley Charity.

Dr. Efthimiou was supported by the Swiss National Science Foundation (Ambizione grant number 180083).

Dr. Mulsant holds and receives support from the Labatt Family Chair in Biology of Depression in Late-Life Adults at the University of Toronto. He also receives support from several grants from the US National Institute of Mental Health and the US Patient-Centered Outcomes Research Institute.

Professor Cipriani is supported by the National Institute for Health Research (NIHR) Oxford Cognitive Health Clinical Research Facility, by an NIHR Research Professorship (grant RP-2017-08-ST2-006), by the NIHR Oxford and Thames Valley Applied Research Collaboration and by the NIHR Oxford Health Biomedical Research Centre (grant BRC-1215-20005). Professor Howes is supported by the Medical Research Council-UK (grant no. MC_A656_5QD30_2135), Maudsley Charity (grant no. 666), and Wellcome Trust (grant no. 094849/Z/10/Z) and the NIHR Biomedical Research Centre at South London and Maudsley NHS Foundation Trust and King’s College London.

The funders had no role in study design, data collection, data analysis, data interpretation, or writing of the report. *The views expressed are those of the author(s) and not necessarily those of H Lundbeck A/s, the NHS/NIHR or the Department of Health*.

