## supplementary for "Reappraising Treatment Effect Heterogeneity in Schizophrenia: A Meta-analysis"

**Supplementary Information**

**Reappraising Variability of Treatment Effects in Schizophrenia: A Meta-analysis**

*General linear model to estimate treatment effects*

We fit a linear model of the following form in the IPD from the RCT:

$$y_{T,i}=\alpha+\beta_{sex} sex_{i}+\beta_{age} age_{i}+\beta_{base} y_{0,i}+\left( \gamma_{sex} sex_{i}+\gamma_{age} age_{i}+\gamma_{base} y_{0,i} \right) treat_{i}+\delta treat_{i}+e_{i}$$

$$e_{i}\sim N(0, s^{2})$$

where $y_{T,i}$ is the final outcome (symptoms at endpoint) for patient $i$, $y_{0,i}$ is baseline symptoms, $sex_{i}$ is 0/1 for male/female, $treat_{i}$ is the 0/1 for placebo/drug.

This model was fit to all patients included in the trial and treatment effects were then estimated for each subject as:

$$\gamma_{sex} sex_{i}+\gamma_{age} age_{i}+\gamma_{base} y_{0,i}+\delta$$

The outcomes in placebo (required for the calculation of $\rho$) were taken as observed in the data, i.e. we only used patients randomized to placebo.

In other words, the treatment condition during the double-blind period was a predictor variable, with sex, age and baseline symptoms as additional predictors. Then based on this model, we estimated outcome for the participants who received placebo had they received treatment (by setting the condition to 1).

*Estimating treatment effect variability*

Previous analyses of variability did not formally estimate treatment effect heterogeneity but instead reported a lack of evidence for heterogeneity on the basis of a non-significant difference between active and placebo effect variabilities, that is a variability ratio (VR) that is not significantly different from zero.

*VR* is defined as follows, with *AT* referring to the active treatment arm and *PL* referring to the placebo treatment arm:

$$VR=\frac{\sigma_{AT}}{\sigma_{PL}}$$

The variable of clinical interest, however, is the variability of the treatment effect ($\sigma_{TE}$). This can be estimated if VR and the correlation ($\rho$) between placebo-effect and treatment-effect are known. For instances where VR≥1, there is a single value of $\frac{\sigma_{TE}}{\sigma_{PL}}$ for any value of $\rho$ :

$$\sigma_{TE}=\sigma_{PL}\left( \sqrt{{VR}^{2}-1+\rho^{2}}-\rho\right)$$

Whereas for instances where VR≤1 there are two possible solutions:

$$\sigma_{TE}=\sigma_{PL}\left( \pm\sqrt{{VR}^{2}-1+\rho^{2}}-\rho\right)$$

In what follows we use the former formula; see some discussion below regarding the second.

After estimating the value for $\rho$ as described in the main paper (i.e. using three different methods, and picking the most conservative estimate for $\rho$) we then calculated $\sigma_{TE}$ for all studies from a recent meta-analysis^1^ given the formula above. The sampling variance for the natural logarithm of $\sigma_{TE}$, $\ln\sigma_{TE}$ was estimated numerically. More specifically, based on previous work, for every study we can estimate the population standard deviation and the sampling variance of $\ln\sigma_{PL}$ as^2^

$\ln\hat{\sigma_{PL}}=\ln s_{PL}+\frac{1}{2(n_{PL}-1)}$ and $S_{\ln\hat{\sigma_{PL}}}^{2}$= $\frac{1}{2(n_{PL}-1)}$

where $s_{PL}$ is the standard deviation of the placebo group, and $n_{PL}$ the number of patients in placebo in that study. Likewise, for the active treatment we have:

$\ln\hat{\sigma_{AT}}=\ln s_{AT}+\frac{1}{2(n_{AT}-1)}$ and $S_{\ln\hat{\sigma_{AT}}}^{2}$= $\frac{1}{2(n_{AT}-1)}$

We now rewrite

$$\sigma_{TE}=\left( \sqrt{{\sigma_{AT}}^{2}-\sigma_{PL}^{2}+\sigma_{PL}^{2}\rho^{2}}-\rho\right)$$

Thus, we need three estimates ($\hat{\sigma_{PL}}$ ,$\hat{\sigma_{AT}}$, $\hat{\rho}$) to obtain an estimate $\hat{\sigma_{TE}}$. Each of these three numbers is estimated in a different way, and they come with their own uncertainty. For $\rho$, we assumed this was a fixed value, namely the most conservative value for the coefficient (but see sensitivity analyses described below; results did not materially change when we used the alternative values for $\rho$).

Next, we want to quantify the uncertainty associated with our estimated $\hat{\sigma_{TE}}$. Unfortunately, there is no simple way to do this in an analytical manner. Instead, we used a sampling method: we generated 10,000 values of $\sigma_{PL}$ and $\sigma_{AT}$ by drawing from their corresponding distributions. For instance, we sampled 10,000 values of $\ln\sigma_{PL}$ by drawing from the normal distribution $N\left( \ln s_{PL}+\frac{1}{2\left( n_{PL}-1 \right)}, \frac{1}{2\left( n_{PL}-1 \right)} \right)$; likewise for $\ln\sigma_{AT}$. For each of these 10,000 pairs we calculated the corresponding $\sigma_{TE}$; finally, from 10,000 $\sigma_{TE}$ we calculated the corresponding standard deviation. This quantifies our uncertainty (standard error) around the estimated value of $\hat{\sigma_{TE}}$. In a sensitivity analysis we also used the uncertainty around the estimation of $\hat{\rho}$ to generate 10,000 values; results regarding the standard errors of $\hat{\sigma_{TE}}$ did not materially change.

Having estimated $\ln\sigma_{TE}$ and the corresponding standard error from each study, we then combined results in a random effects meta-analysis (note that the results presented in text are as $\sigma_{TE}$ rather than its logarithm, for interpretability). To centre the $\sigma_{TE}$ around a clinically meaningful value we also calculated an estimate of the mean treatment effect by performing a random effects meta-analysis of unadjusted mean difference between placebo and active treatment. At this point readers should note that the estimated $\ln\sigma_{TE}$ across studies are correlated, owing to the fact that they are all calculated using the same values for $\rho$. Our analysis disregards this correlation, and this could in principle be a limitation, leading to narrower confidence intervals. However, we calculated the correlations induced across the study-specific estimates and we found them to be very small for most cases. Thus, disregarding these induced correlations is expected to have minimal impact on our results.

In some trials the observed VR was not compatible with our main estimate of $\rho$ (in cases of low VR a greater absolute value of $\rho$ is required for solutions to contain only real numbers). These trials were removed in our main analysis. In a sensitivity analysis we used the most liberal value for the correlation coefficient obtained from the three methods described in the paper. Our results showed an even more pronounced variability of treatment effects , specifically the standard deviations for total, positive, and negative symptoms were 23.3, 7.4, and 3.7 points respectively.

For studies in which VR<1 there exists an additional second solution:

$$\sigma_{TE}=\sigma_{PL}\left( -\sqrt{{VR}^{2}-1+\rho^{2}}-\rho\right)$$

We presented only the findings for the first solution:

$$\sigma_{TE}=\sigma_{PL}\left( \sqrt{{VR}^{2}-1+\rho^{2}}-\rho\right)$$

This is because the alternative of using the second solution for those studies with VR<1 leads to a situation in which there is a large discontinuities for the estimate of $\sigma_{TE}$ between a study with a VR of e.g. 1.01 and a study with a VR of e.g. 0.99. This discontinuity appears implausible, and we have therefore restricted the possible solutions.

Screening

Included

Eligibility

Identification

Additional records identified through other sources
(n = 73)

Records after duplicates removed
(n =7,936)

Records screened
(n = 7,936)

Records excluded
(n =6,805)

Full-text articles assessed for eligibility
(n = 1,131 )

Full-text articles excluded (n = 1,067)

Studies included in qualitative synthesis
(n = 66)

Studies included in quantitative synthesis (meta-analysis)
(n = 66)

Records identified through database searching
(n = 11, 006 )

PRISMA Flow Diagram

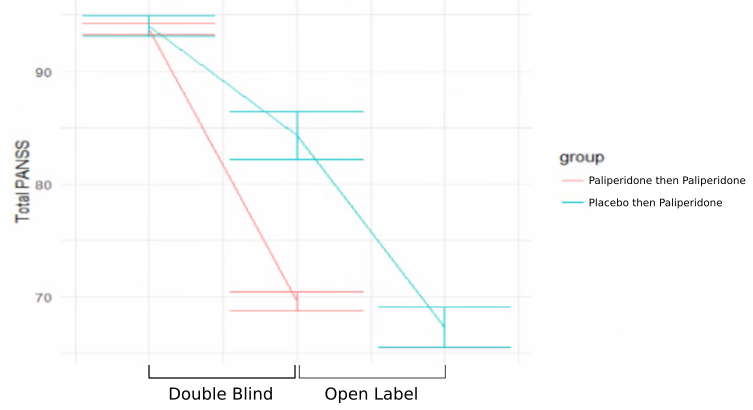

**eFigure 1**

Mean PANSS score at the end of the double blind period for patients that received active paliperidone (69.5, SD 19) was very similar to the mean PANSS scores at the end of the open label period for patients that initially received placebo (67.2, SD 19.9) (Independent sample t- test t=0.93, p = 0.34). This observation provides support for the the assumptions behind the “open label” method described in our paper.

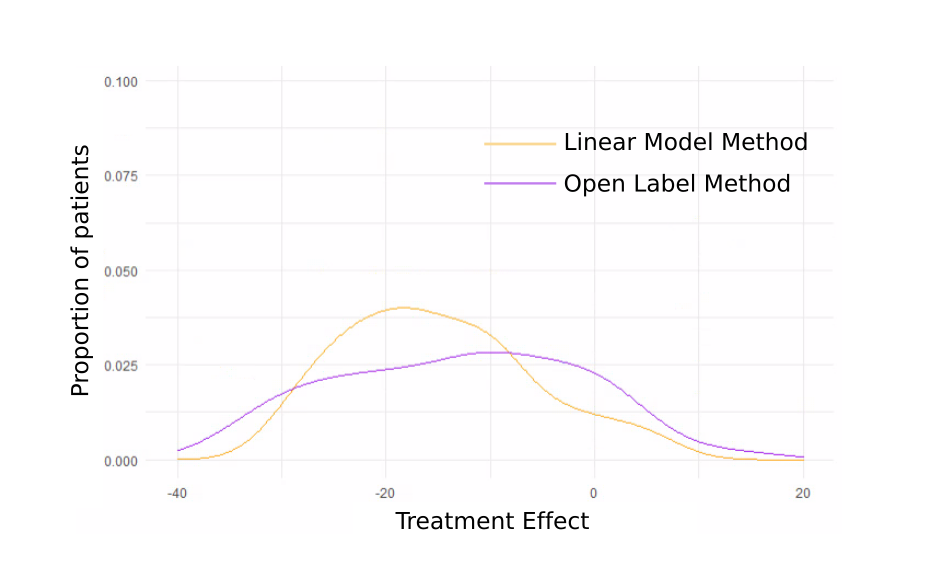

**eFigure 2**

Density plot of treatment effects. Both methods of individual treatment effect estimation show clinically significant inter-individual differences.

### PRISMA Checklist

| **Section/topic** | **#** | **Checklist item** | **Reported on page #** |
| --- | --- | --- | --- |
| **TITLE** | | |  |
| Title | 1 | Identify the report as a systematic review, meta-analysis, or both. | 1 |
| **ABSTRACT** | | |  |
| Structured summary | 2 | Provide a structured summary including, as applicable: background; objectives; data sources; study eligibility criteria, participants, and interventions; study appraisal and synthesis methods; results; limitations; conclusions and implications of key findings; systematic review registration number. | 3 |
| **INTRODUCTION** | | |  |
| Rationale | 3 | Describe the rationale for the review in the context of what is already known. | 3-5 |
| Objectives | 4 | Provide an explicit statement of questions being addressed with reference to participants, interventions, comparisons, outcomes, and study design (PICOS). | 5 |
| **METHODS** | | |  |
| Protocol and registration | 5 | Indicate if a review protocol exists, if and where it can be accessed (e.g., Web address), and, if available, provide registration information including registration number. | 6 |
| Eligibility criteria | 6 | Specify study characteristics (e.g., PICOS, length of follow-up) and report characteristics (e.g., years considered, language, publication status) used as criteria for eligibility, giving rationale. | 6 |
| Information sources | 7 | Describe all information sources (e.g., databases with dates of coverage, contact with study authors to identify additional studies) in the search and date last searched. | 6-7 |
| Search | 8 | Present full electronic search strategy for at least one database, including any limits used, such that it could be repeated. | 6, supplement |
| Study selection | 9 | State the process for selecting studies (i.e., screening, eligibility, included in systematic review, and, if applicable, included in the meta-analysis). | 6-7 |
| Data collection process | 10 | Describe method of data extraction from reports (e.g., piloted forms, independently, in duplicate) and any processes for obtaining and confirming data from investigators. | 6-7 |
| Data items | 11 | List and define all variables for which data were sought (e.g., PICOS, funding sources) and any assumptions and simplifications made. | 6-7 |
| Risk of bias in individual studies | 12 | Describe methods used for assessing risk of bias of individual studies (including specification of whether this was done at the study or outcome level), and how this information is to be used in any data synthesis. | 7 |
| Summary measures | 13 | State the principal summary measures (e.g., risk ratio, difference in means). | 7-8 |
| Synthesis of results | 14 | Describe the methods of handling data and combining results of studies, if done, including measures of consistency (e.g., I^2^) for each meta-analysis. | 8-9 |

| **Section/topic** | **#** | **Checklist item** | **Reported on page #** |
| --- | --- | --- | --- |
| Risk of bias across studies | 15 | Specify any assessment of risk of bias that may affect the cumulative evidence (e.g., publication bias, selective reporting within studies). | 7-10 |
| Additional analyses | 16 | Describe methods of additional analyses (e.g., sensitivity or subgroup analyses, meta-regression), if done, indicating which were pre-specified. | 7-9 |
| **RESULTS** | | |  |
| Study selection | 17 | Give numbers of studies screened, assessed for eligibility, and included in the review, with reasons for exclusions at each stage, ideally with a flow diagram. | 11, supplement |
| Study characteristics | 18 | For each study, present characteristics for which data were extracted (e.g., study size, PICOS, follow-up period) and provide the citations. | supplement |
| Risk of bias within studies | 19 | Present data on risk of bias of each study and, if available, any outcome level assessment (see item 12). |  |
| Results of individual studies | 20 | For all outcomes considered (benefits or harms), present, for each study: (a) simple summary data for each intervention group (b) effect estimates and confidence intervals, ideally with a forest plot. | 11-12 |
| Synthesis of results | 21 | Present results of each meta-analysis done, including confidence intervals and measures of consistency. | 11-12 |
| Risk of bias across studies | 22 | Present results of any assessment of risk of bias across studies (see Item 15). | 12 |
| Additional analysis | 23 | Give results of additional analyses, if done (e.g., sensitivity or subgroup analyses, meta-regression [see Item 16]). | 12-14 |
| **DISCUSSION** | | |  |
| Summary of evidence | 24 | Summarize the main findings including the strength of evidence for each main outcome; consider their relevance to key groups (e.g., healthcare providers, users, and policy makers). | 2,3,15 |
| Limitations | 25 | Discuss limitations at study and outcome level (e.g., risk of bias), and at review-level (e.g., incomplete retrieval of identified research, reporting bias). | 16-17 |
| Conclusions | 26 | Provide a general interpretation of the results in the context of other evidence, and implications for future research. | 2,3,17 |
| **FUNDING** | | |  |
| Funding | 27 | Describe sources of funding for the systematic review and other support (e.g., supply of data); role of funders for the systematic review. | 18 |
